# Clonal Hematopoiesis and Incident Solid Cancers Resolved by Histologic Subtype: A Prospective Cohort Study

**DOI:** 10.64898/2026.11.05.26367360

**Authors:** Parham Habibzadeh, Kun Zhao, Alexis Cenname, Yu Jen Alexander Jan, Emily Geramita, Annie Im, John D. Sorkin, Alexander G. Bick, Dennis J. Hsu

## Abstract

Clonal hematopoiesis of indeterminate potential (CHIP) is a recognized precursor of myeloid malignancy and an independent cardiovascular risk factor, but its relationship with solid tumors is incompletely defined, particularly at histologic resolution. Here, we quantify the association between CHIP and incident cancer across hematologic and solid-organ sites, by clone size, mutation number, and histologic subtype in a prospective cohort of UK Biobank participants with baseline genetic sequencing and cancer ascertained by national registry linkage; those with pre-baseline cancer were excluded. Vanderbilt BioVU was used for replication. CHIP was associated with any incident cancer (HR, 1.22; 95% CI, 1.18-1.27) and with all solid tumors combined (HR, 1.13; 95% CI, 1.08-1.19). Among 17 endpoints, two survived multiple testing correction: lung adenocarcinoma (HR, 1.79; 95% CI, 1.48-2.16) and esophageal squamous cell carcinoma (HR, 2.28; 95% CI, 1.37-3.81). Risk increased with clone size and multi-hit CHIP, most markedly for esophageal cancer (VAF ≥30%: HR, 2.72), and the esophageal squamous association persisted in 5-year landmark analysis (HR, 2.57). In BioVU, CHIP was directionally associated with esophageal cancer and lung adenocarcinoma and its association with all solid tumors overall (HR, 1.24; 95% CI, 1.17-1.32) was confirmed. These findings extend CHIP from a hematologic and cardiovascular risk marker to a risk marker of solid-organ cancer risk with histologic resolution and motivate clone-size–aware risk models and mechanistic studies.

## Introduction

Clonal hematopoiesis of indeterminate potential (CHIP) describes the outgrowth of a hematopoietic stem cell clone carrying a leukemia-associated somatic mutation in a person who has neither a dysplastic hematopoiesis nor an unexplained cytopenia.^1^ Felt largely to be a phenomenon of aging, CHIP is detectable in a few percent of adults in midlife but increased in prevalence to include over 10% of those older than 70 years, with mutations in *DNMT3A*, *TET2*, and *ASXL1* genes accounting for most driver events.^2–5^ As a result of large sequencing efforts, the genetic architecture of clonal hematopoiesis and inherited factors that favor clonal expansion have been largely identified.^6,7^ Furthermore, longitudinal studies have established CHIP as a stepwise precursor from which myeloid neoplasms arise.^8^

The clinical implications of CHIP extend well beyond the bone marrow. Carriers face a higher risk of atherosclerotic cardiovascular disease, and the responsible mutant myeloid cells appear to act through uncontrolled inflammatory pathways.^9–11^ Whether the same biology raises the risk of solid tumors has been harder to definitively establish. CHIP is common among patients with non- hematologic cancers, partly because cytotoxic and radiation therapy select for mutant clones,^12,13^ which complicates inference from cross-sectional oncology studies. Tissue-level research has shown that clones infiltrate solid tumors (*i.e.,* tumor-infiltrating clonal hematopoiesis) and that their presence worsens outcomes in lung and other cancers.^14^ Furthermore, a nested case-control study linked clonal hematopoiesis to incident lung cancer.^15^ Even so, prospective population-scale cohort-derived evidence that CHIP precedes and predicts solid-organ cancer, resolved by histologic subtype and by features of the clone itself, remains sparse.^16^

We therefore used the UK Biobank, a prospective cohort, to examine whether individuals with CHIP go on to develop solid tumors at higher rates compared to those without and, if so, which cancers account for the excess risk. CHIP carriers and noncarriers, none with cancer at baseline, were compared in cohorts matched on the propensity to carry CHIP and further adjusted for shared risk factors including age and smoking status, with a landmark analysis to reduce the influence of preclinical disease. In addition, since tumors grouped under a single organ can differ in their cell of origin and risk factors,^17^ the lung and esophageal cancers were analyzed by histologic subtypes. We then sought to replicate the principal findings in an independent US biobank, Vanderbilt BioVU.

## Methods

### Study Populations and Design

The discovery cohort comprised UK Biobank participants with whole-exome sequencing of blood collected at the baseline assessment (2006-2010). Herein, the exposure was measured on enrollment. Incident cancers were identified through linkage to national cancer and death registries linked to the UK Biobank through January 2024. Participants with any cancer diagnosed before baseline were excluded. We required complete data on core covariates (age, sex, Townsend deprivation index, and body mass index [BMI]) and positive follow-up time. Our final cohort available for propensity score matching was 409,150 participants (**Figure 1**). Replication analyses were performed in Vanderbilt BioVU, a de-identified electronic health record (EHR)-linked biobank at Vanderbilt University Medical Center. The BioVU cohort included 245,540 individuals. After excluding participants with a prior hematologic malignancy (n = 8,491), 237,049 individuals remained. Participants younger than 40 years of age were further excluded, yielding a final analytic cohort of 139,685 individuals.

**Figure 1.**
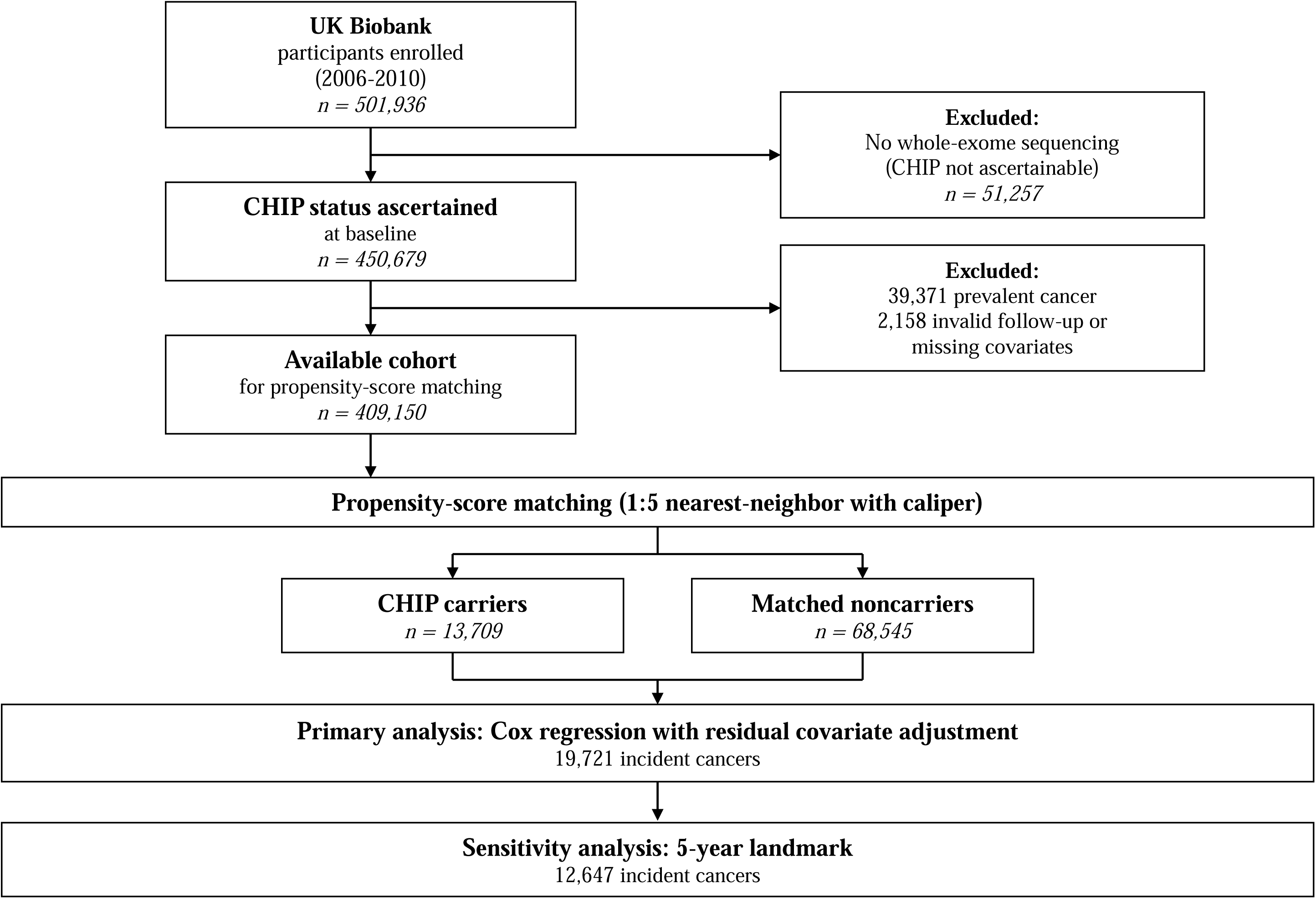
Study Discovery Cohort Analytic Design. Flow of UK Biobank participants from enrollment to the analysis. CHIP carriers were matched 1:5 to noncarriers using propensity score and compared using Cox proportional hazards regression, with a 5-year landmark analysis performed as a sensitivity analysis. CHIP, clonal hematopoiesis of indeterminate potential.

### Ascertainment of CHIP

CHIP was defined as a somatic mutation in a canonical myeloid driver gene present at a variant allele fraction of 2% or greater in the absence of a hematologic malignancy.^1,18^ Variants were called from exome data and curated with a published framework that combines sequence- and population-level filtering to limit germline and artifactual variants.^18^ Beyond a binary exposure (present vs. absent), we characterized CHIP in three ways that capture distinct aspects of clonal biology: (1) by individual driver gene; (2) by clone size, using the maximum variant allele fraction to define small (<10%), expanded (≥10%), large (≥20%), and very large (≥30%) clones; and (3) by mutation number, contrasting a single mutation with multi-hit CHIP (≥2 mutations).

### Outcomes

The primary outcome was any incident cancer. Cancers were assigned to anatomic sites using ICD-10 codes and, for the lung and esophagus, were further classified by histology from International Classification of Diseases for Oncology (ICD-O) morphology codes into adenocarcinoma and squamous cell carcinoma, with small-cell and other non–small-cell categories for the lung. Person-time was measured from baseline to the earliest of cancer diagnosis, death, loss to follow-up, or the end of national registry follow up. In BioVU, histologic subtype was determined using the Vanderbilt Tumor Registry based on ICD-O morphology codes, with esophageal squamous cell carcinoma defined by squamous histology. For the cancer phenome-wide analysis, cancer phenotypes were defined using validated phecodes mapped from ICD-9 and ICD-10 diagnosis codes.

### Covariates

All models were adjusted for age, sex, Townsend deprivation index, self-reported ethnicity, smoking status (never, former, current), regular alcohol use (yes vs. no), BMI (kg/m^2^), diabetes (yes vs. no), chronic kidney disease (yes vs. no), and family history of cancer (yes vs. no). These variables were selected because they are shared determinants of both clonal expansion and the cancers under study and felt to be the most plausible confounders. In BioVU, models adjusted for age, sex, smoking status, BMI, hypertension, diabetes mellitus, and the first five genetic principal components.

### Statistical Analysis

In the discovery cohort, we estimated hazard ratios (HRs) using Cox proportional hazards regression within propensity score–matched cohorts. Propensity scores were generated using a logistic-regression model where the outcome variable was the CHIP status which was regressed over the covariates above. Each carrier was matched with up to five noncarriers by nearest neighbor on the logit of the propensity score.^19^

We further screened the association between the binary CHIP exposure (variant allele fraction ≥2%) and 17 prespecified cancer endpoints, comprising three lung cancer histologies (adenocarcinoma, squamous cell carcinoma, and small cell carcinoma), two esophageal cancer histologies (adenocarcinoma and squamous cell carcinoma), and site-level endpoints for colorectal, breast, prostate, melanoma, pancreatic, gastric, kidney, bladder, liver, ovarian, uterine, and thyroid cancer. In subsequent exploratory analyses, we further examined the association by driver gene, clone size (variant allele fraction strata), and mutation number (single vs ≥2). Furthermore, a 5-year landmark analysis excluding the events and follow-up during the first 5 years after blood draw was carried out. Tests were 2-sided, and both Benjamini-Hochberg and Bonferroni-adjusted P values were reported for the 17-endpoint screen. Statistical analysis was done using Python language and the associated libraries (lifelines and scikit-learn).^20,21^

For the BioVU replication cohort, associations between CHIP and esophageal cancer outcomes were evaluated using multivariable Cox proportional hazards regression among incident cases, adjusting for age, sex, smoking status, body mass index (BMI), hypertension, diabetes mellitus, and the first five genetic principal components. A cancer-focused association study was additionally performed to evaluate associations between CHIP and site-specific cancer phecodes using the same covariate adjustment.

### Ethics

The Vanderbilt University Medical Center Institutional Review Board approved the BioVU cohort study. The UK Biobank holds ethical approval from the North West Multi-centre Research Ethics Committee, and all participants gave written informed consent (application No. 709086). Reporting follows the STROBE guideline for cohort studies.

## Results

### Study population

Of 409,150 UK Biobank participants who met the inclusion criteria, 13,709 (3.4%) carried CHIP at baseline. During the follow up period, 77,677 incident cancers occurred, of which 39,160 were solid tumors. Propensity score matching produced a cohort of 82,254 participants (13,709 carriers and 68,545 matched noncarriers) in which all covariates were balanced (maximum absolute standardized mean difference, 0.007) with a median follow up of 14.4 (IQR, 11.8-15.3) years in the matched cohort. The crude incidence rate of any cancer in the matched cohort was 22.3 versus 18.2 per 1000 person-years in carriers versus noncarriers (**Table 1**).

**Table 1.**
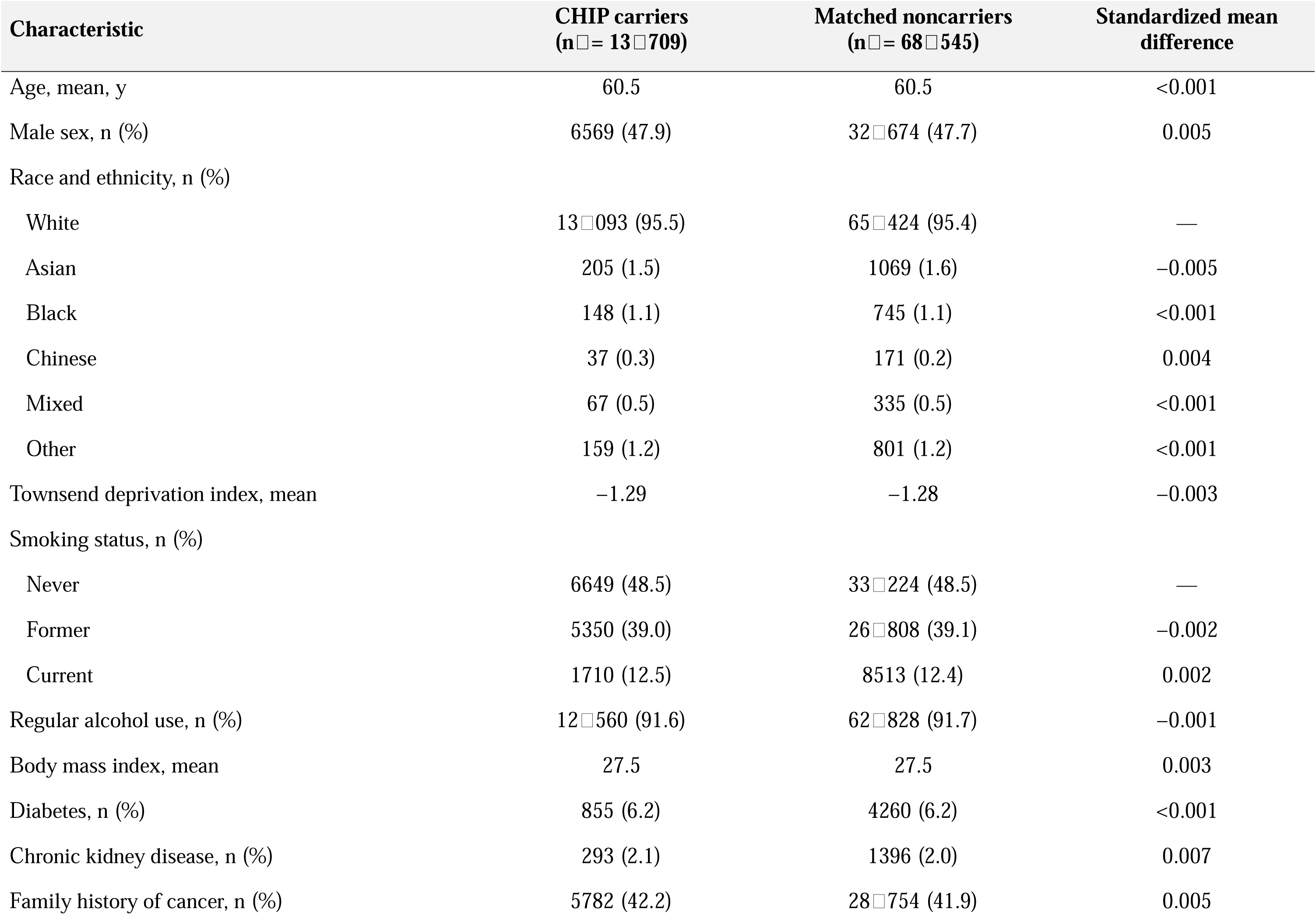
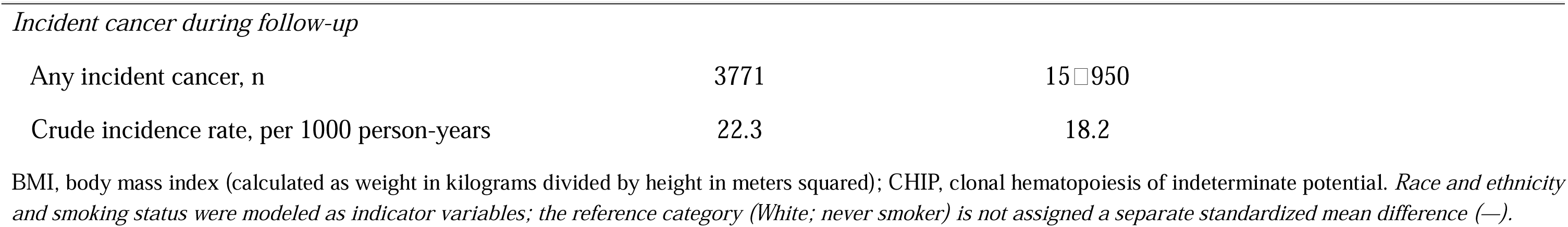
Baseline Characteristics of the Propensity Score-Matched Cohort.

| **Characteristic** | **CHIP carriers<br>(n□= 13□709)** | **Matched noncarriers<br>(n□= 68□545)** | **Standardized mean<br>difference** |
|---|---|---|---|
| Age, mean, y | 60.5 | 60.5 | <0.001 |
| Male sex, n (%) | 6569 (47.9) | 32□674 (47.7) | 0.005 |
| Race and ethnicity, n (%) |  |  |  |
| White | 13□093 (95.5) | 65□424 (95.4) | — |
| Asian | 205 (1.5) | 1069 (1.6) | −0.005 |
| Black | 148 (1.1) | 745 (1.1) | <0.001 |
| Chinese | 37 (0.3) | 171 (0.2) | 0.004 |
| Mixed | 67 (0.5) | 335 (0.5) | <0.001 |
| Other | 159 (1.2) | 801 (1.2) | <0.001 |
| Townsend deprivation index, mean | −1.29 | −1.28 | −0.003 |
| Smoking status, n (%) |  |  |  |
| Never | 6649 (48.5) | 33□224 (48.5) | — |
| Former | 5350 (39.0) | 26□808 (39.1) | −0.002 |
| Current | 1710 (12.5) | 8513 (12.4) | 0.002 |
| Regular alcohol use, n (%) | 12□560 (91.6) | 62□828 (91.7) | −0.001 |
| Body mass index, mean | 27.5 | 27.5 | 0.003 |
| Diabetes, n (%) | 855 (6.2) | 4260 (6.2) | <0.001 |
| Chronic kidney disease, n (%) | 293 (2.1) | 1396 (2.0) | 0.007 |
| Family history of cancer, n (%) | 5782 (42.2) | 28□754 (41.9) | 0.005 |
| *Incident cancer during follow-up* |  |  |  |
| Any incident cancer, n | 3771 | 15□950 |  |
| Crude incidence rate, per 1000 person-years | 22.3 | 18.2 |  |

BMI, body mass index (calculated as weight in kilograms divided by height in meters squared); CHIP, clonal hematopoiesis of indeterminate potential. *Race and ethnicity and smoking status were modeled as indicator variables; the reference category (White; never smoker) is not assigned a separate standardized mean difference (—).*

### CHIP and the overall burden of incident cancer

CHIP was associated with any incident cancer (HR, 1.22; 95% CI, 1.18-1.27) and with all solid tumors combined (HR, 1.13; 95% CI, 1.08-1.19). As expected, the association with hematologic malignancy was far stronger (all hematologic: HR, 3.07; 95% CI, 2.76-3.40; myeloid neoplasms: HR, 10.49; 95% CI, 8.63-12.74). Estimates were essentially identical in unmatched covariate-adjusted models (any incident cancer: HR, 1.22; 95% CI, 1.18-1.26).

### Histology-resolved screen across 17 prespecified endpoints

In the prespecified screen of the binary CHIP exposure against 17 cancer endpoints (histology-resolved for lung and esophagus, site-level otherwise), two endpoints passed both Benjamini-Hochberg false-discovery-rate control and the more conservative Bonferroni threshold: lung adenocarcinoma (HR, 1.79; 95% CI, 1.48-2.16) and esophageal squamous cell carcinoma (HR, 2.28; 95% CI, 1.37-3.81; FDR q = 0.01; Bonferroni P = 0.03) (**Figure 2**). Several additional endpoints reached nominal significance but did not survive correction, including prostate, bladder, and ovarian cancer (HRs, 1.11, 1.33, and 1.42, respectively). No association was observed for colorectal, breast, pancreatic, renal, gastric, hepatic, uterine, or thyroid cancer or for melanoma.

**Figure 2.**
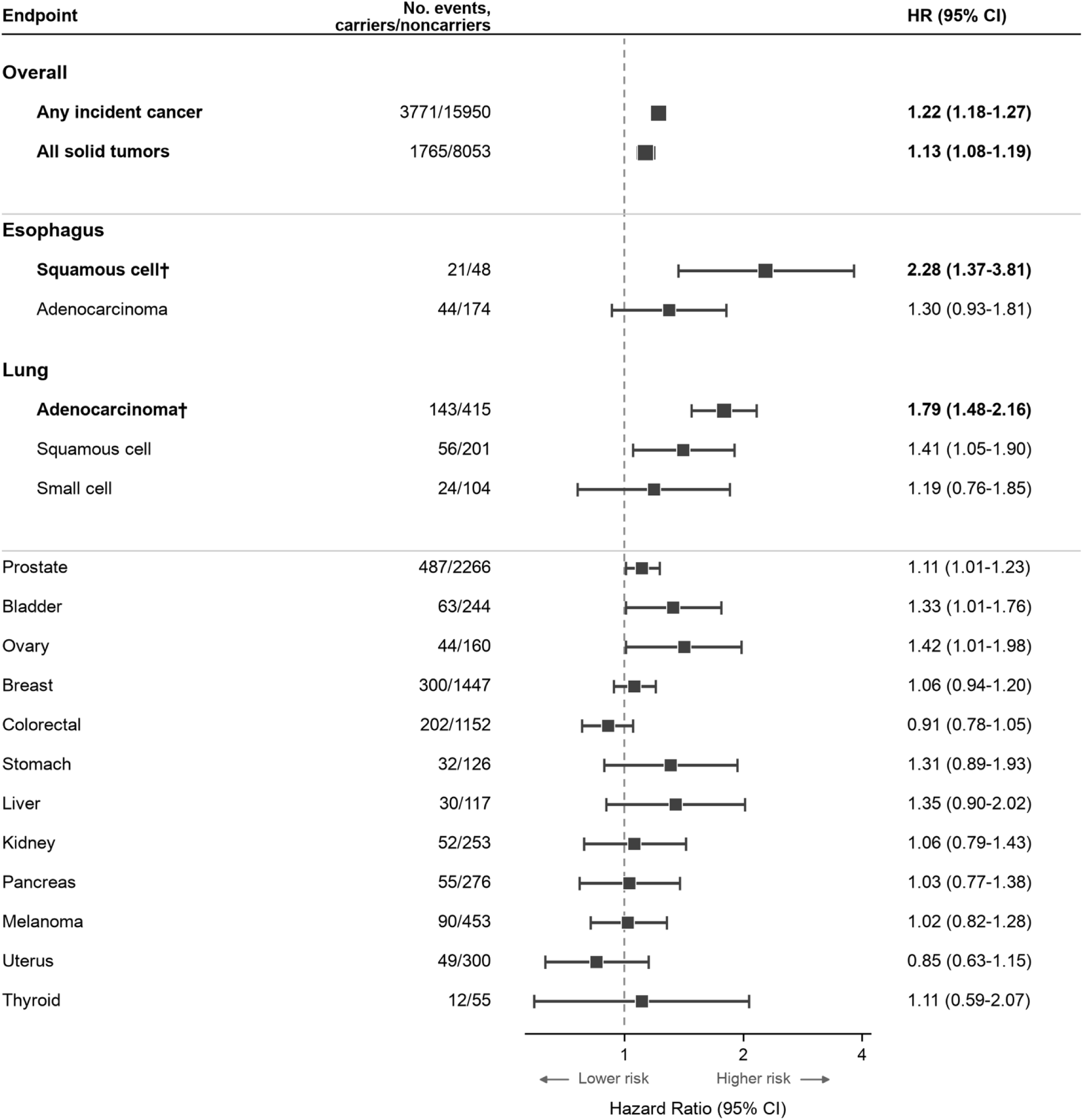
Association of CHIP With Incident Cancer in the UK Biobank. Hazard ratios (squares) and 95% CIs (horizontal lines) from Cox proportional hazards regression in the 1:5 propensity score–matched cohort. Estimates marked with a dagger (†), esophageal squamous cell carcinoma and lung adenocarcinoma, passed both Benjamini-Hochberg and Bonferroni correction. The dashed line marks a hazard ratio of 1; the axis is logarithmic. CHIP indicates clonal hematopoiesis of indeterminate potential; CI, confidence interval; HR, hazard ratio.

The value of histologic resolution was most apparent in esophageal tumors. The organ-level association was driven by the squamous subtype, whereas esophageal adenocarcinoma showed a weaker association that did not pass a priori defined thresholds for statistical significance. A parallel pattern was seen in the lung, where the adenocarcinoma association exceeded that for the squamous and small cell subtypes (**Figure 2**).

### Clone size and mutational complexity exploratory analyses

For all solid tumors combined, small clones (variant allele fraction [VAF] <10%) conferred no excess risk (HR, 1.01; 95% CI, 0.92-1.10), whereas risk emerged at a VAF of 10% or greater and increased to an HR of 1.23 (95% CI, 1.12-1.35) at a VAF of 20% or greater, plateauing at 1.21 (95% CI, 1.05-1.39) for the largest clones (VAF ≥ 30%). For esophageal cancer the gradient was steeper, reaching an HR of 2.72 (95% CI, 1.39-5.30) at a VAF of 30% or greater; within the squamous subtype specifically, large clones (VAF ≥ 20%) carried an HR of 3.03 (95% CI, 1.12-8.21). The highest VAF stratum contained too few squamous events for stable estimation.

Mutational complexity showed the same direction of effect. Multi-hit CHIP (≥2 mutations) was associated with all solid tumors combined (HR, 1.38; 95% CI, 1.13-1.69) and, more strongly, with esophageal cancer (HR, 3.33; 95% CI, 1.45-7.66) (**Figure 3**). Among individual drivers, DNMT3A CHIP was associated with esophageal squamous cell carcinoma (HR, 1.96; 95% CI, 1.06-3.63); TET2 yielded too few squamous events for subtype-specific estimation but showed a positive association with esophageal cancer at the organ level (HR, 1.95; 95% CI, 1.01-3.79).

**Figure 3.**
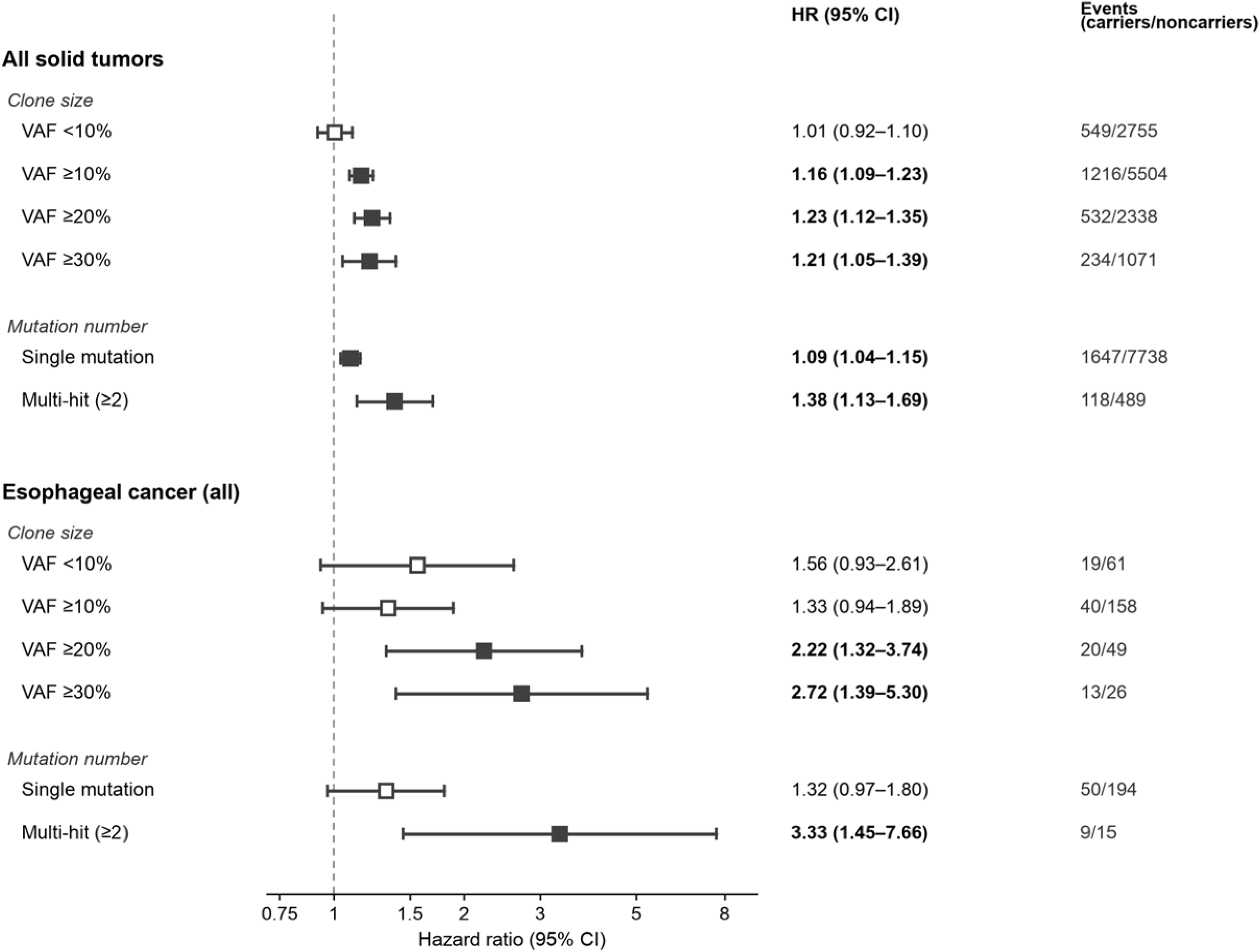
CHIP and Cancer Risk by Clone Size and Mutational Complexity in the UK Biobank. Hazard ratios (squares) and 95% CIs (lines) for CHIP carriers versus 1:5 propensity score–matched noncarriers, for all solid tumors combined and esophageal cancer, shown by clone size and mutation number. Filled squares indicate a 95% CI excluding 1. The dashed line marks a hazard ratio of 1; the axis is logarithmic. CHIP, clonal hematopoiesis of indeterminate potential; CI, confidence interval; HR, hazard ratio; VAF, variant allele fraction.

### Landmark analysis

To address reverse causation, we repeated the primary analyses after excluding events and follow-up during the first 5 years after blood draw. The associations were preserved or strengthened: any cancer (HR, 1.24; 95% CI, 1.19-1.30), all solid tumors (HR, 1.14; 95% CI, 1.07-1.22), lung adenocarcinoma (HR, 1.95; 95% CI, 1.54-2.47), and esophageal squamous cell carcinoma (HR, 2.57; 95% CI, 1.44-4.61).

### Replication in Vanderbilt BioVU

In the BioVU biobank, CHIP was associated with solid tumors overall (HR, 1.24; 95% CI, 1.17-1.32) similar to the estimate from the discovery cohort for all solid tumors.

We were able to perform a histology-matched replication in the BioVU among 48 incident cases of lung adenocarcinoma and found that CHIP was directionally concordant with the discovery association (HR, 1.30; 95% CI, 0.96-1.76). For esophageal cancer specifically, CHIP was associated with risk in the same direction as in the discovery cohort, although the estimate did not reach significance (covariate-adjusted HR, 1.22; 95% CI, 0.85-1.75 among 34 incident cases). A histology-specific replication of the squamous finding was not feasible: although 82 of 978 esophageal cancers in BioVU were squamous, only 1 incident squamous cell carcinoma occurred among CHIP carriers, and therefore a reliable estimate could not be obtained.

## Discussion

In a prospective cohort of more than 400,000 adults with blood sampled years before cancer diagnosis, CHIP was associated with a modest increase in overall solid-tumor risk and, most prominently, with esophageal squamous cell carcinoma, one of only two histology-resolved endpoints to survive both false-discovery-rate and Bonferroni correction across a 17-endpoint screen. Risk scaled with clone size and was amplified in multi-hit carriers across solid tumors and at the esophageal organ level, and the esophageal squamous association persisted in landmark analysis that excluded the first 5 years of follow-up. In an independent US biobank of about 250,000 total participants, CHIP was associated with solid tumors overall and was directionally concordant for esophageal cancer at the organ level.

These findings extend the clinical significance of CHIP beyond its established roles in myeloid malignancy^8^ and atherosclerotic cardiovascular disease^11^ to the risk of selected solid tumors. Overall, these associations are consistent with, and refine, prior observations. A nested case-control study previously linked clonal hematopoiesis to incident lung cancer;^15^ our data confirm that association at population scale and localize it predominantly to the adenocarcinoma subtype. A prospective analysis of the Women’s Health Initiative linked CHIP to solid-tumor incidence but with a site-specific pattern, an association with breast, though not lung or colorectal, cancer, illustrating that CHIP–solid-tumor relationships are heterogeneous across sites and underscoring the value of histologic resolution.^22^ Furthermore, recent work has shown that mutant clones infiltrate solid tumors and worsen outcomes,^14^ and a high prevalence of CHIP among patients with non-hematologic cancers has been reported in multiple cross-sectional series, in part reflecting therapy-induced selection.^12,13^ However, although prior studies have linked CHIP to the broader solid-tumor risk, a prospective, histology-resolved investigation especially for esophageal cancer, has not, to our knowledge, been previously described.

In the current study, confounding by shared risk factors, age and smoking status in particular, was addressed by propensity score matching with additional covariate adjustment in the outcome model.^19^ In addition, misclassification of the exposure, in which an undiagnosed tumor sheds mutant DNA that is mistaken for CHIP, is unlikely because CHIP was ascertained from blood collected in 2006-2010, years before the cancers were diagnosed, and the 2% variant-allele-fraction floor limits contamination by circulating tumor DNA.^18^ Reverse causation is likewise unlikely as the association between CHIP and esophageal squamous carcinoma persisted, and actually strengthened in the landmark analysis (*i.e.,* when the first 5 years of follow-up were excluded). The clone-size gradient and the multi-hit association further suggest a biologically graded relationship rather than an artifact.

A plausible biological explanation for these associations is the pro-inflammatory phenotype of mutant myeloid cells. Loss-of-function TET2 mutations impair the resolution of inflammation and increase interleukin-6–driven signaling,^10^ and TET2-deficient clones amplify NLRP3-inflammasome–mediated interleukin-1β signaling and accelerate inflammatory disease in experimental models,^23^ mechanisms that have been proposed to connect aging and inflammation more broadly.^9^ These same cytokines are directly implicated in esophageal squamous carcinogenesis. In a transgenic mouse model, overexpression of interleukin-1β in the squamous epithelium was by itself sufficient to drive the stepwise progression from hyperplasia through dysplasia to invasive squamous cell carcinoma, accompanied by recruitment of myeloid cells and elevated interleukin-6 and tumor necrosis factor–α; notably, this progression occurred even under germ-free conditions, isolating inflammation rather than microbial exposure as a proximate driver of squamous cell carcinomas.^24^ In clinical samples, interleukin-1β is correspondingly overexpressed in human esophageal squamous cell carcinoma, where it promotes tumor growth and invasion.^25^ These observations support the possibility that individuals harboring CHIP may be predisposed to esophageal squamous cell cancer via aberrant interleukin signaling, however this hypothesis is not testable by the present epidemiologic data and would require additional mechanistic and tissue-based studies to confirm.

The divergence between esophageal squamous cell carcinoma and adenocarcinoma also carries a methodologic lesson. The two tumors differ in cell of origin, risk-factor profile, and geographic distribution and are distinct at the molecular level. Esophageal squamous cell carcinoma resembles squamous carcinomas of other organs, whereas esophageal adenocarcinoma is more similar to the chromosomally unstable variant of gastric adenocarcinoma.^17,26^ Therefore, collapsing them into a single organ-level endpoint dilutes a subtype-specific signal; the organ-level HR of 1.52, understates the squamous-specific HR of 2.28. This has direct implications for replication. In BioVU, as in most Western populations, esophageal cancer is predominantly adenocarcinoma,^17^ so the organ-level replication estimate is weighted toward the subtype for which CHIP shows no discovery-cohort association; the modest, nonsignificant organ-level HR in BioVU is not surprising. BioVU thus supports the pan-solid-tumor and organ-level associations while remaining directionally consistent with, but underpowered for, the squamous-specific finding. The scarcity of incident squamous cases in BioVU precluded a histology-matched replication and a histology-resolved meta-analysis, which we regard as an important direction for confirmation in cohorts enriched for squamous cell carcinoma.

This study has limitations. Although the discovery cohort is large, the number of incident esophageal squamous events were relatively rare (69 cases), and subtype-, gene-, and VAF-stratified estimates are correspondingly imprecise, as reflected in the wide HR confidence intervals. The UK Biobank is predominantly of European ancestry and is subject to healthy-volunteer selection, which may limit generalizability, particularly to populations in which esophageal squamous cell carcinoma is most common. Replication was limited to a single US biobank and, for the squamous endpoint specifically, was underpowered. Finally, as an observational study, these data establish association rather than causation. Nonetheless, several features strengthen confidence in these findings, including propensity-score matching with close covariate balance, a histology-resolved design, an exploratory dose-response relationship with clone size, amplified risk in multi-hit carriers, persistence in a five-year landmark analysis, and directional replication in an independent biobank.

In conclusion, our findings position CHIP as a systemic risk marker that extends into solid-organ oncology and motivate clone-size-aware risk assessment and direct mechanistic study of the role of clonal hematopoiesis in epithelial carcinogenesis.

## Data availability

Individual-level data from both cohorts are subject to controlled access and cannot be redistributed by the study authors to protect participant privacy. UK Biobank data were analyzed under Application No. 709086 and are available to researchers through UK Biobank (https://www.ukbiobank.ac.uk/register-apply/). For Vanderbilt BioVU, deidentified patient data is available to researchers whose proposed use of the data has been approved with a signed data access agreement by contacting.

## Author Contributions

P.H. designed the research, performed statistical analysis, analyzed and interpreted data, and wrote the manuscript; K.Z. performed the BioVU analyses and analyzed and interpreted data, and wrote the manuscript; A.C. assisted with statistical analysis and validation; Y.J.A.J., E.G., and A.I. interpreted data and critically revised the manuscript; J.D.S. advised on statistical analysis and critically revised the manuscript; A.G.B. contributed data and analytical tools for the BioVU biobank and critically revised the manuscript; D.J.H. designed the research, supervised the study, interpreted data, and critically revised the manuscript; and all authors approved the final manuscript.

## Declaration of generative AI and AI-assisted technologies in the manuscript preparation process

During the preparation of this work, the authors used Anthropic Claude Opus for language editing and for suggesting alternative phrasings. The authors reviewed and edited the output as needed and take full responsibility for the content of the published article.

